# An evaluation of the i-gel Plus® supraglottic airway device in elective patients: the interim results from a prospective international multicentre study

**DOI:** 10.1101/2024.02.16.24302948

**Authors:** J. Werner, O. Klementova, Jan Bruthans, Jaromir Macoun, Tomasz Gaszynski, Tomas Henlin, Will Donaldson, Erik Lichnovsky, Shiva Arava, Ana M Lopez, Raquel Berge, Pavel Michalek

**Author notes:** Correspondence to: P. Michalek. these authors contributed equally (J. Werner, O. Klementova).

## Abstract

The i-gel Plus supraglottic airway represents the next generation of the i-gel device. The aim of this international multicentre prospective cohort study was to evaluate its performance in adult patients during elective procedures in various surgical disciplines under general anaesthesia. The primary outcome of the study was the overall success rate of insertion allowing effective airway management, ventilation, and oxygenation. The secondary outcomes included perioperative performance of the device and the incidence of postoperative adverse events. The data of the first 1000 patients from our study (455 males and 545 females) are presented. These patients were mostly operated on in the supine position (83.3%) with a minority of them being in the lateral or lithotomy positions. The overall success rate was 98.6%, with a first-attempt success rate of insertion of 88.1%. A significant difference between males and females was seen for the overall success rate – 97.4% vs. 99.6% (p=0.002) but not for the successful insertion on the first attempt (p=0.97) The mean oropharyngeal seal pressure was 32 (±7) cmH_2_O. The only independent factor increasing the risk of first-attempt failure was low experience of the operator (p<0.001). The insertion of the device was rated by 80.3% as being either very easy or easy. Fibreoptic assessment through the i-gel Plus showed a full view of the vocal cords in 67.8% of patients, a partial view in 21.9% and a downfolded epiglottis in 9.4% of patients. A gastric tube was inserted in 11.2% of patients with a 99.1% success rate. Perioperative complications included desaturation below 85% in 0.6%, traces of blood on the device in 7.4%, laryngospasm in 0.5% and gastric contents inside the cuff in 0.2% of patients. There were no clinical signs of aspiration and a 0.1% incidence of bronchospasm. Severe postoperative sore throat was recorded in 1.4%, and long-term hoarse voice in 0.2% of patients. All patients with moderate and serious postoperative complaints are being followed up by phone at 3 and 6 months. The i-gel Plus seems to be an effective supraglottic airway device providing a high success rate of insertion, sufficient oropharyngeal seal pressure, and a reasonably low incidence of complications.

**Trial registration:** ISRCTN86233693

## Introduction

Supraglottic airway devices have played an important role in elective anaesthetic practice and airway management for more than three decades [1]. They are preferred as less invasive alternatives to tracheal intubation in fasted patients and for procedures outside of the abdominal and thoracic cavities [2]. Since the invention of the first laryngeal mask airway in the 1980s, supraglottic airway devices have been significantly developed to offer high quality performance and a low incidence of perioperative and postoperative adverse effects [3]. Various supraglottic airway devices with differing mechanisms of seal have been introduced into clinical anaesthesia within the last two decades.

The i-gel Plus® supraglottic airway device (Intersurgical Ltd., Wokingham, United Kingdom) is a modified version of the i-gel® supraglottic airway, which was introduced into clinical practice in 2007 [4]. The cuff of the device is made from a soft thermoplastic elastomer, with a non-inflatable bowl, which is anatomically preshaped in order to mirror the vocal cords and perilaryngeal structures. Compared to the previous version of the device, the i-gel Plus® has a number of different features which should theoretically improve its performance (Fig. 1).

**Fig. 1.**
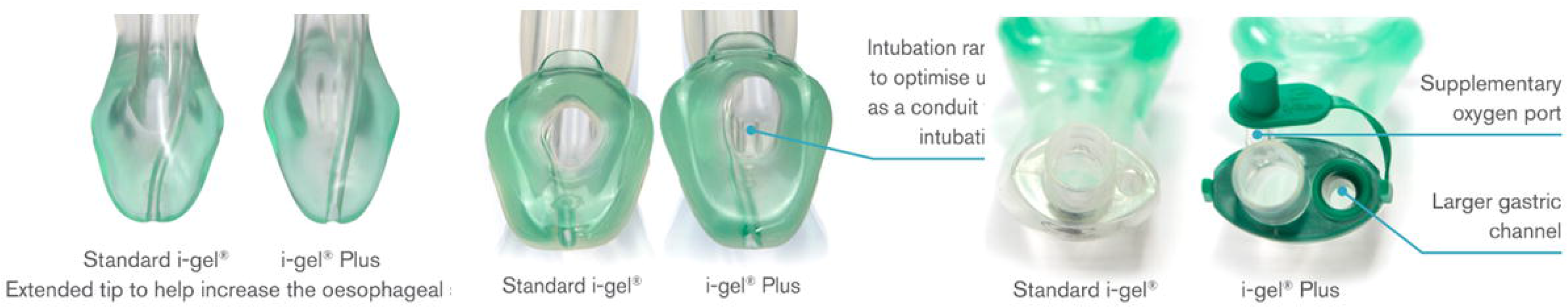
Comparison of the design of the i-gel and i-gel Plus supraglottic airway devices

They include a wider diameter of the gastric drainage channel, a ramp located at the bottom of the ventilation channel (which should improve the success rate of blind intubation through the device), a slight prolongation of the tip for improvement of the oesophageal seal and a supplementary oxygen port.

This study aims to evaluate the clinical performance, perioperative and postoperative complications of the i-gel Plus® supraglottic airway device in a multicentre multinational prospective study. The study aims to enrol at least 2000 adult patients scheduled for elective procedures under general anaesthesia. In this manuscript, we would like to present preliminary results based on the initial 1000 patients. With the primary outcome being the total success rate of insertion we hypothesized that at least a 95 % success rate is required for this device to be considered effective.

## Methods

The study protocol (version 1.2, released October 1st, 2019) was prepared in full concordance with the Helsinki Declaration and subsequently approved by the ethics and research committees of all seven hospitals taking part in the study (Appendix 1). The study was registered prior to enrolment of the first patient with the ISRCTN Clinical Trial Registry (ISRCTN86233693, December 16th, 2019). The full protocol including data collection/management, plan for statistical analysis, oversight, storage, security and dissemination of data was published [5]. The protocol and reporting of data have been in adherence with the Strengthening the Reporting of Observational Studies in Epidemiology (STROBE) statement [6]. All participants received a Patient Information Sheet at least 24 h in advance, had an opportunity to discuss any aspect of the study before enrollment with the study anaesthetist, and signed an informed consent form. By December 1st, 2023 a total of 1000 patients had been enrolled. Whilst all patient materials were in the relevant language for each centre (English, Czech, Spanish, Polish), the CRF was only in English. The paper forms were transferred into an electronic database (www.redcap.vfn.cz). Three members of the study team performed a check of the electronic forms for completeness and accuracy on a weekly basis.

Inclusion criteria were: ASA class I-III, both sexes, age 18-89 years, elective surgery with planned airway management using a supraglottic airway (SGA) device, whilst exclusion criteria involved acute procedures, non-fasted patients, any increased risk for aspiration of gastric contents, body mass index (BMI) higher than 35 kg.m^-2^, prone, sitting or steep head down patient positionings, shared airway or intracavity surgeries, and inability to understand or sign informed consent.

Preoperatively, demographic parameters and difficult airway assessment predictive values (Mallampati class, thyromental distance, neck extension, mandible protrusion and neck circumference) were recorded.

The study flow chart is provided in Fig. 2. Patients were fasted for 6 h from solid food and 2 h from clear liquids. They received alprazolam 0.5 mg or midazolam 7.5 mg orally 1 h before induction of general anaesthesia. Intravenous induction of anaesthesia was performed using propofol and an opioid (sufentanil, fentanyl, alfentanil, remifentanil) in standard doses. The administration of muscle relaxants was not part of the protocol, however, if required for any reason during the surgery, it was to be reported in detail to the CRF. The properly lubricated i-gel Plus device was inserted after loss of the eyelash reflex. Anaesthesia was maintained according to the operator’s preference – inhalational or TIVA with opioids. Minimum monitoring included pulse oximetry, non-invasive blood pressure measurement, ECG, and capnography. The choice of device size was performed based on the manufacturer’s recommendation with the actual body weight for patients with BMI ≤ 25 kg.m^-2^ and with the adjusted body weight for patients with BMI 25-35 kg.m^-2^.

**Fig. 2.**
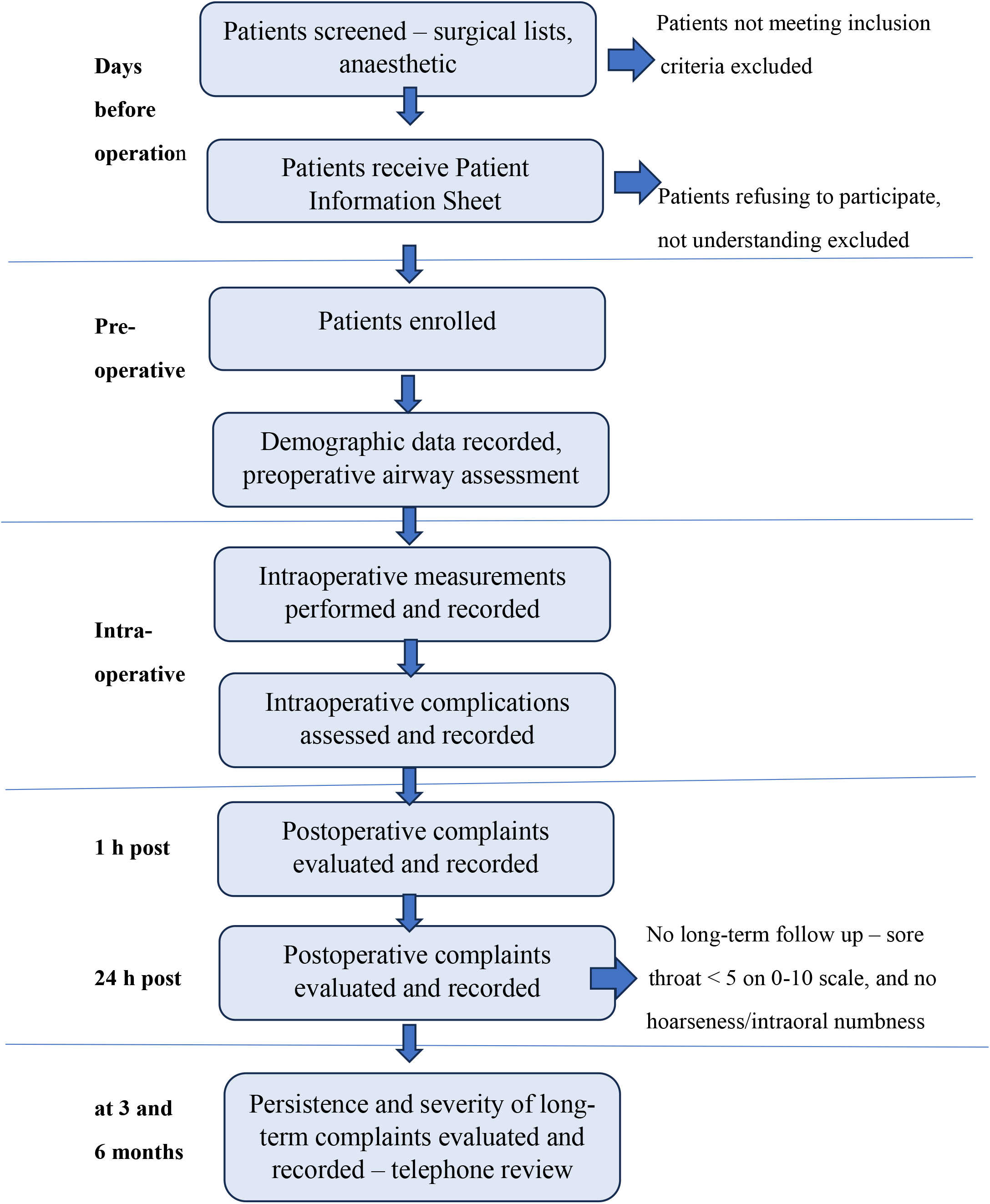
Study flowchart

The primary outcome was total success rate which was defined as, the ability to deliver sufficient tidal volumes until the end of surgery without any audible leak around the i-gel Plus. The secondary outcomes included: intraoperative measures, intraoperative complications, and early and late postoperative patient complaints. Intraoperative secondary outcomes measured were: first attempt insertion success rate, number of insertion attempts, insertion time, lowest saturation during induction on pulse oximetry, seal pressure, necessity of adjustment manoeuvers during the procedures, difficulty of insertion as reported by the operators, fiberoptic evaluation of the position of the vocal cords through the device (optional) and ease of gastric tube insertion through the i-gel Plus (optional). The fiberoptic evaluation was only performed when a flexible bronchoscope was available and the gastric tube was inserted when clinically indicated, mainly in patients with signs of a distended stomach, in laparoscopies or for prolonged procedures. Recorded intraoperative complications were: detection of blood or gastric contents on the i-gel Plus after removal and clinical presentation of aspiration of gastric contents - laryngospasm or bronchospasm. Postoperative complaints were assessed at 1 h and 24 h post procedures. Those assessed were: sore throat, difficulty in swallowing, hoarseness, feeling of numbness inside the oral cavity, neck pain, and jaw pain. All were evaluated on a 0-10 verbal scale. Those patients reporting sore throat, pain during swallowing, neck or jaw pain with a score of more than 5 on the scale, and all those with any hoarseness or numbness inside the oral cavity at 24 hours were followed up by telephone review at both 3 and 6 months.

### Statistics

Sample size calculation was performed prior to the creation of the study protocol using a modified Cochran formula [7]. Based on the results of other large studies evaluating SGAs we set up a 95% overall success rate with a 95% CI 93-97%. The minimum required sample size was 1924 patients, and the plan was to enroll 2000 participants to compensate for dropouts.

A statistical plan for the trial was already set up in the protocol [7]. It involved a comparison between male and female patients, a comparison between different sizes of the devices, and a comparison between the performance of operators of different seniority. A manual logistic multivariable regression was performed in order to find the factors influencing total and first-attempt failures. The R Core Team (version 2023, https://www.R-project.org) was used for all statistical measurements. P-values of less than 0.05 were deemed as statistically significant. In cases with a low number of observations the Fisher exact test was used for comparisons (primary outcome). Pearson’s chi-squared test was performed for other comparisons of success rates. Differences in seal pressure between the sexes and sizes of the device were evaluated by Welch’s two-sample t-test.

## Results

In total, data from 1000 patients enrolled between 23/09/2020 and 29/12/2023 were analyzed. There were 545 females (54.5%) and 455 (45.5%) males in this cohort. Their demographic data, type of surgical specialty, and patient positioning are provided in Table 1. The most commonly used size of the device was a 4, which was inserted in 569 (56.9%) patients, in both males and females. Size 3 was used in 149 (14.9%) only in female patients, while a size 5 was inserted in 282 (28.2%), and predominantly in male patients.

**Table 1.** Demographic parameters and characteristic of the procedures.

|  | Mean [95% CI] or total number (%) |
| --- | --- |
| Sex | 455 (45.5%) males<br>545 (54.5%) females |
| Age (years) | 52.7 [51.7-53.7], range 18-88 |
| BMI (kg.m-2) | 26.4 [26.1-26.7], range 15.5-35 |
| ASA class I-III | 376 (37.6%)/517 (51.7%)/107 (10.7%) |
| Mallampati class I-IV | 422 (42.2%)/464 (46.4%)/105 (10.5%)/<br>9 (0.9%) |
| Thyromental distance (cm) | 7.2 [7.1-7.4], range 2.5-14 |
| Mandibular protrusion (good/limited/poor) | 821 (82.1%)/176 (17.6%)/3 (0.3%) |
| Neck circumference (cm) | 39.6 [39.2-39.9], range 18-60 |
| Cervical spine mobility (normal/limited/none) | 941 (94.1%)/58 (5.8%)/1(0.1%) |
| Mouth opening (cm) | 4.6 [4.6-4.7], range 2-9 |
| Dentition (sound/crowns,bridges,venirs/toothless, dentures) | 678 (67.8%)/218 (21.8%)/104(10.4%) |
| Beard, moustache | 115 (11.5%) |
| Procedure type | General surgery: 388 (38.8%)<br>Orthopaedics/Trauma: 326 (32.6%)<br>Urology: 103 (10.3%)<br>Gynaecology: 98 (9.8%)<br>Vascular surgery: 59 (5.9%)<br>Other: 26 (2.6%) |
| Patient positioning | Supine: 831 (83.1%)<br>Gynaecological: 154 (15.4%)<br>Head down: 2 (0.2%)<br>Head up, semi-sitting: 2 (0.2%)<br>Lateral: 9 (0.9%)<br>Other: 2 (0.2%) |
| Anaesthesia duration (min) | 61.4 [59.2-63.5], range 7-260 |
| Type of anaesthesia | General with volatiles: 910 (91%)<br>Total intravenous: 90 (9.0%)<br>Muscle relaxants used: 156 (15.6%) |

Detailed data on perioperative device performance and the incidence of postoperative complaints are provided in Table 2. The i-gel Plus provided sufficient ventilation and oxygenation until the end of the procedure in 986 (98.6%) of patients. Failure was recorded in 12 males and 2 females (p = 0.002) which makes a 2% difference of the overal success rate between the sexes. Out of these 14 failed insertions, seven (50%) patients were subsequently successfully managed with another supraglottic airway device (LMA Supreme, LM AuraGain) while the remaining seven (50%) underwent tracheal intubation.

**Table 2.** Perioperative and postoperative parameters.

|  | Mean [95% CI] or number (%) | p value |
| --- | --- | --- |
| Overall insertion success rate | 986 (98.6) |  |
| - Males | 443 (97.4) |  |
| - Females | 543 (99.6) | 0.002 |
| First-attempt success rate | 882 (88.2) |  |
| - Males | 401 (88.1) |  |
| - Females | 481 (88.3) | 0.97 |
| Additional airway manoeuvres | 127 (13) |  |
| - Change of the device position | 92 |  |
| - Head or neck positioning | 64 |  |
| - Jaw thrust | 27 |  |
| Oropharyngeal seal pressure (cmH<sub>2</sub>O) | 31.9 [31.5-32.3] |  |
| - Males | 31.2 [30.8-31.6] |  |
| - Females | 32.4 [32.0-32.8] | 0.01 |
| Insertion time (s) | 13.4 [12.5-14.3] |  |
| Lowest spO<sub>2</sub> during induction (%) | 97 [96.7-97.2] |  |
| Fibreoptic evaluation | 240 (24.3) |  |
| - Vocal cords completely visible | 165 (68.8) |  |
| - Partial view to the vocal cords | 51 (21.3) |  |
| - Only epiglottis visible | 22 (9.2) |  |
| - No clear structures identified | 2 (0.8) |  |
| Gastric tube insertion | 111 (11.3) |  |
| - Very easy | 62 (55.9) |  |
| - Easy | 45 (40.5) |  |
| - Neutral | 3 (2.7) |  |
| - Difficult | 0 |  |
| - Very difficult | 1 (0.9) |  |

The initial attempt of the i-gel Plus insertion was successful in 882 (88.2%) of patients, and did not differ significantly between males and females (p = 1.00). The first attempt success rate was also not different between the sizes of the devices (p = 0.185). However, the insertion on the initial attempt was significantly affected by the seniority of the anaesthesia provider (p < 0.001). Senior doctors working more than 5 years in anaesthesia achieved better first-attempt success rates than less experienced physicians in training. Change of size of the device was necessary in 21 (2.1%) of insertions. Manual regression evaluating patient and operator factors affecting the first-attempt failure rate of insertion revealed that only the anaesthetist’s higher seniority significantly decreased the risk of this event (Table 3).

**Table 3.**
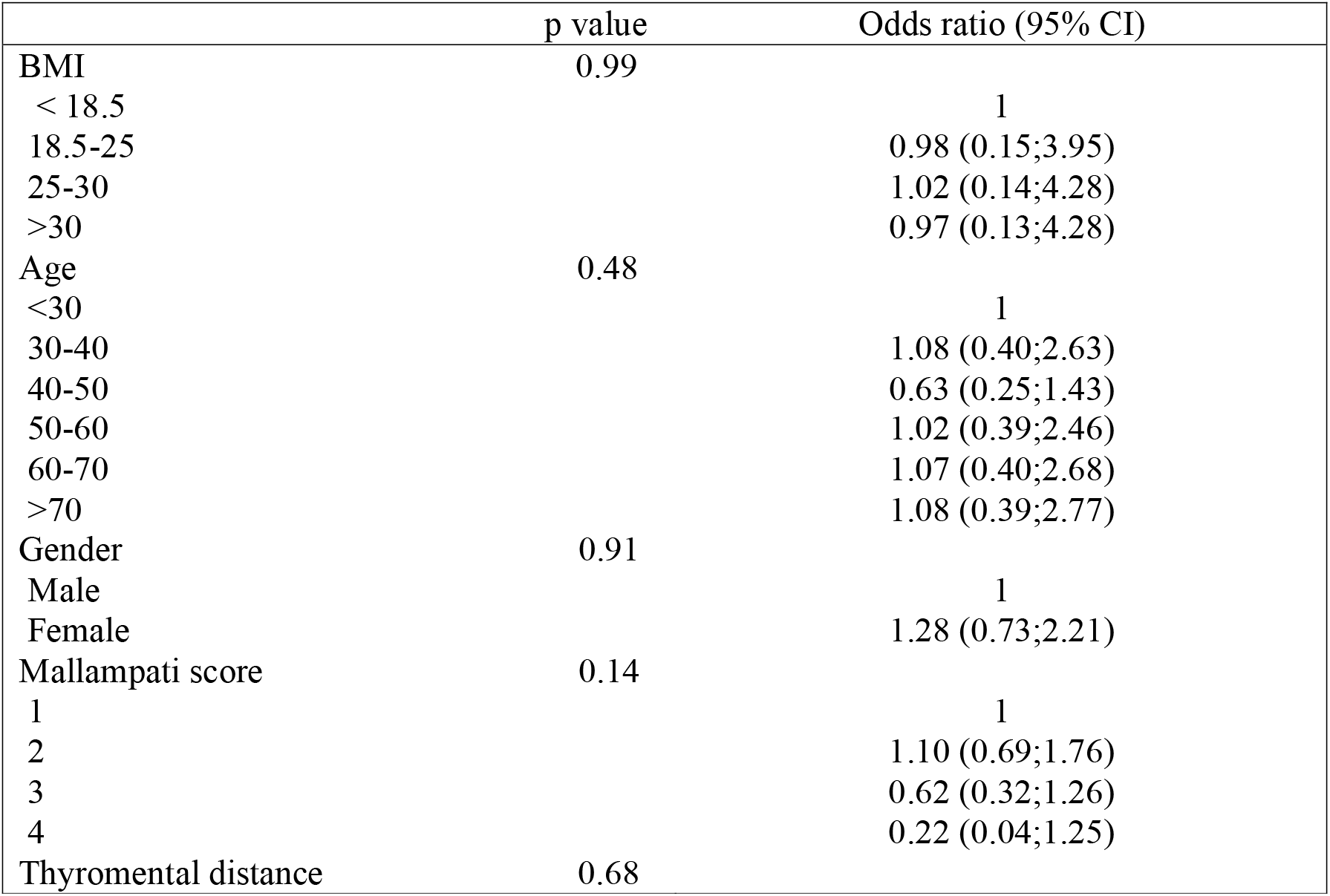
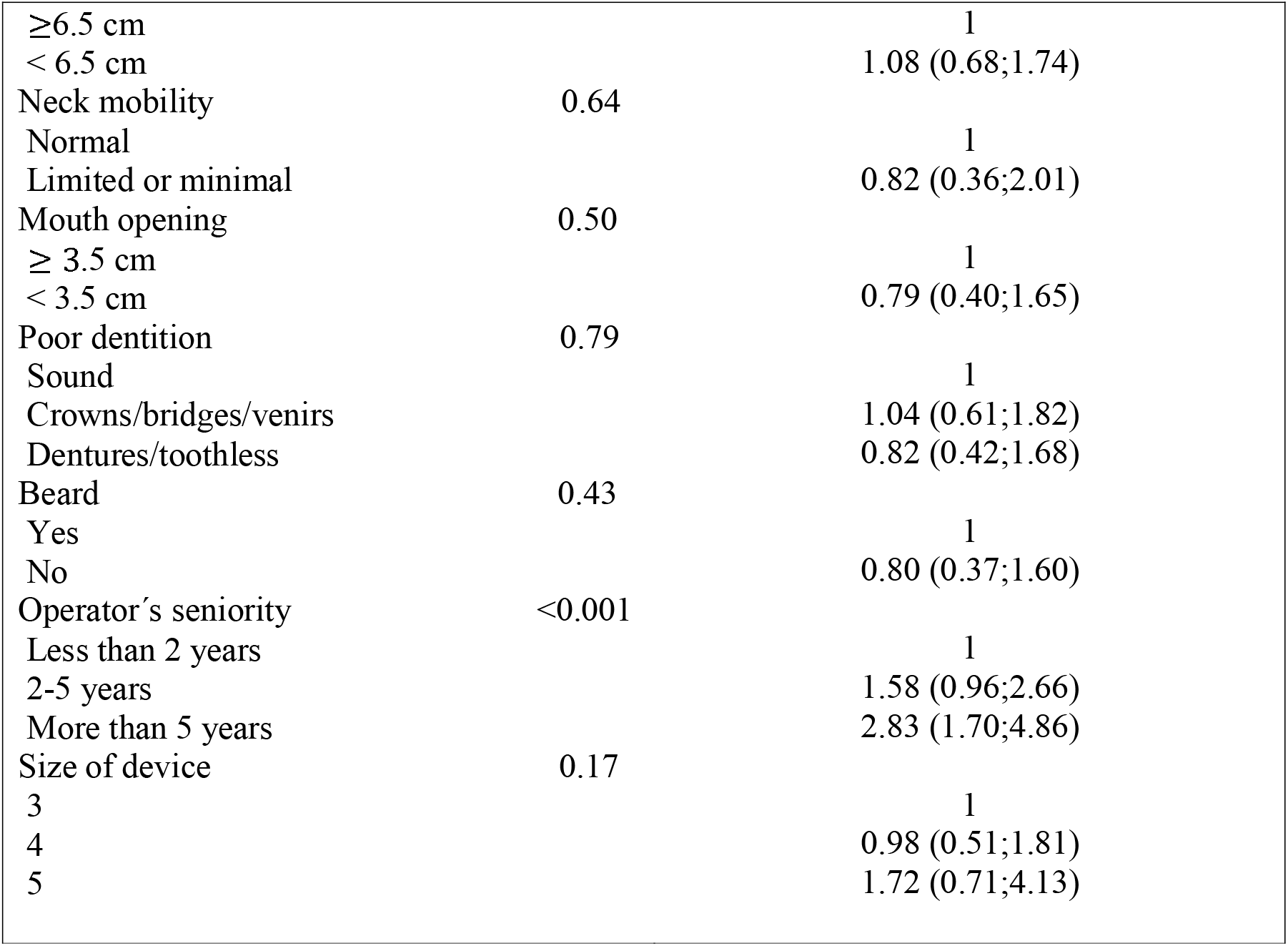
Risk factors for first-attempt insertion failure of the i-gel Plus. Effect size provided as odds ratio for incidencies. CI = confidence interval.

|  | p value | Odds ratio (95% CI) |
| --- | --- | --- |
| BMI | 0.99 |  |
| < 18.5 |  | 1 |
| 18.5-25 |  | 0.98 (0.15;3.95) |
| 25-30 |  | 1.02 (0.14;4.28) |
| >30 |  | 0.97 (0.13;4.28) |
| Age | 0.48 |  |
| <30 |  | 1 |
| 30-40 |  | 1.08 (0.40;2.63) |
| 40-50 |  | 0.63 (0.25;1.43) |
| 50-60 |  | 1.02 (0.39;2.46) |
| 60-70 |  | 1.07 (0.40;2.68) |
| >70 |  | 1.08 (0.39;2.77) |
| Gender | 0.91 |  |
| Male |  | 1 |
| Female |  | 1.28 (0.73;2.21) |
| Mallampati score | 0.14 |  |
| 1 |  | 1 |
| 2 |  | 1.10 (0.69;1.76) |
| 3 |  | 0.62 (0.32;1.26) |
| 4 |  | 0.22 (0.04;1.25) |
| Thyromental distance | 0.68 |  |
| ≥6.5 cm |  | 1 |
| < 6.5 cm |  | 1.08 (0.68;1.74) |
| Neck mobility | 0.64 |  |
| Normal |  | 1 |
| Limited or minimal |  | 0.82 (0.36;2.01) |
| Mouth opening | 0.50 |  |
| ≥ 3.5 cm |  | 1 |
| < 3.5 cm |  | 0.79 (0.40;1.65) |
| Poor dentition | 0.79 |  |
| Sound |  | 1 |
| Crowns/bridges/venirs |  | 1.04 (0.61;1.82) |
| Dentures/toothless |  | 0.82 (0.42;1.68) |
| Beard | 0.43 |  |
| Yes |  | 1 |
| No |  | 0.80 (0.37;1.60) |
| Operator's seniority | <0.001 |  |
| Less than 2 years |  | 1 |
| 2-5 years |  | 1.58 (0.96;2.66) |
| More than 5 years |  | 2.83 (1.70;4.86) |
| Size of device | 0.17 |  |
| 3 |  | 1 |
| 4 |  | 0.98 (0.51;1.81) |
| 5 |  | 1.72 (0.71;4.13) |

The mean oropharyngeal seal pressure after the insertion was 32 (±7) cmH_2_O (Fig. 3). The difference between males and females, although statistically significant, reached only 1 cmH_2_O which is not of clinical importance. Seal pressures of lower than 20 cmH_2_O, where the peak inspiratory pressure may exceed the seal pressure were recorded in 48 (4.9%) patients. On the contrary, a maximum seal pressure of 40 cmH_2_O was achieved in 255 (25.9%) of cases. The mean insertion time was 13.4 (±14.6) sec, with a mean time of achieving effective gas exchange of 19.3 (±16.1) sec. A capnography trace was recorded in 90% of patients within 30 sec after discontinuation of the face-mask ventilation. Intraoperative manipulation of the device, including change of device position, head or neck movements, or application of jaw thrust was required in 127 (13%) patients.

**Fig. 3.**
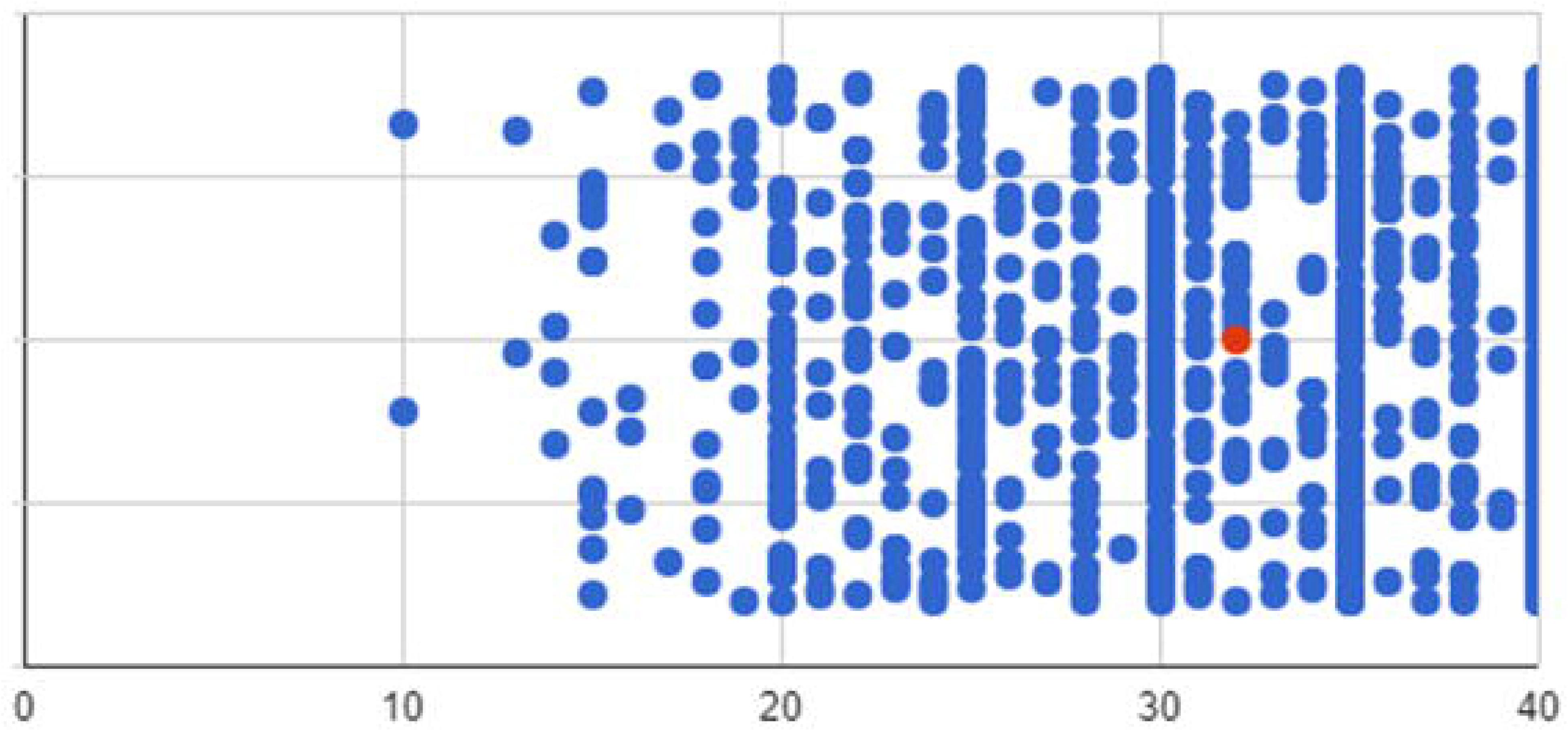
Scatterplot of the oesophageal seal pressures

In total, 240 (24.3%) procedures involved fibreoptic examination through the i-gel Plus. We used the scale published by Kapila et al. [8]. Full or partial view of the vocal cords was recorded in 90.1% of cases (Fig. 4), while the epiglottis was downfolded and obstructing the view in 9.2% of cases, and neither the laryngeal inlet nor epiglottis was visible in 0.2% of insertions, despite effective oxygenation and ventilation. The gastric tube was inserted in 111 (11.4%) of procedures and it was considered very easy or easy in 96.4% of them.

**Fig. 4.**
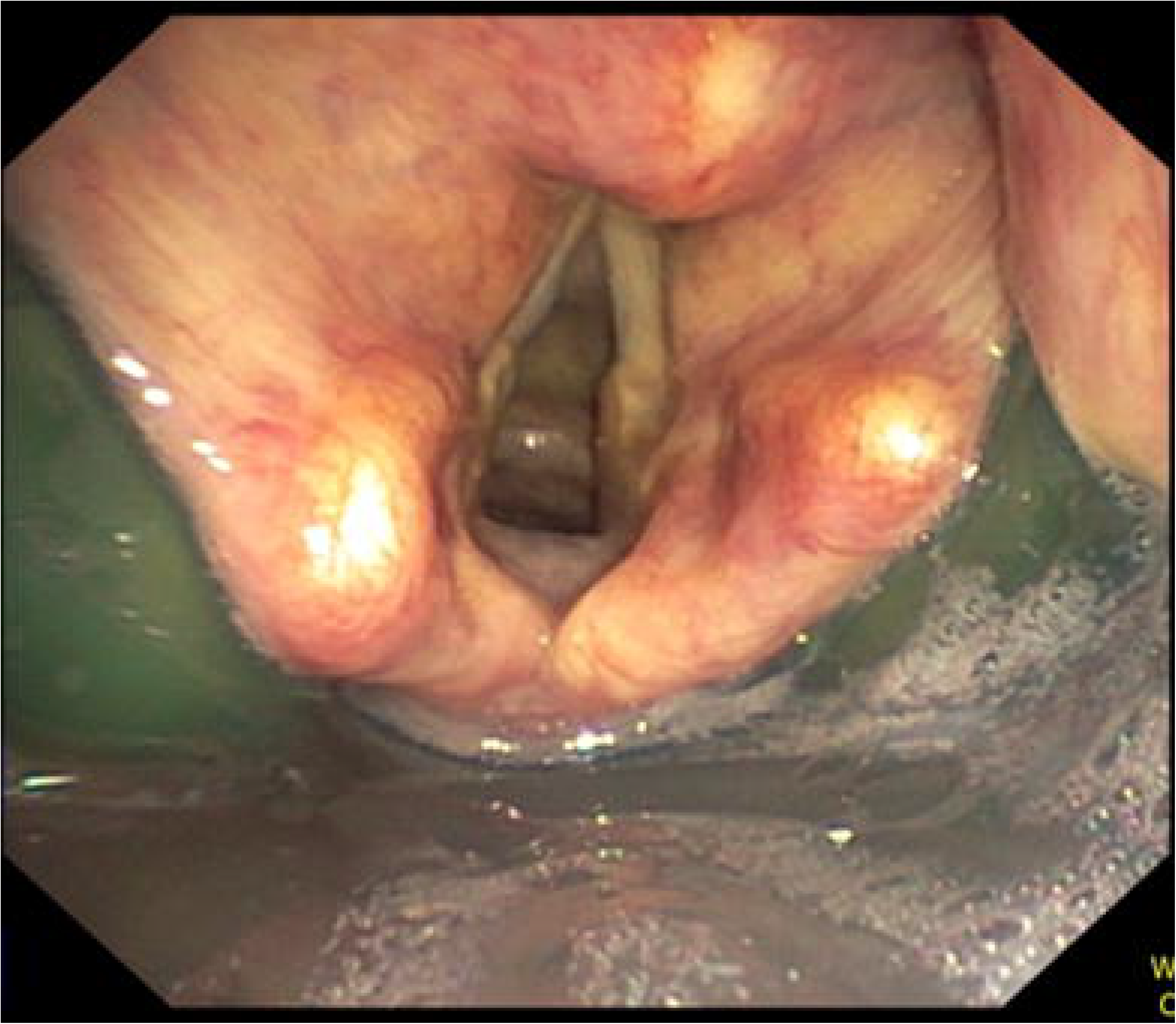
Fibreoptic evaluation through the i-gel Plus showing full view of the vocal cords

The satisfaction with the device insertion and performance was assessed by the operators on a 1-5 Likert scale. Very easy or easy insertion was reported by 80.3% of anaesthetists, neutral by 15.2%, and difficult or very difficult insertion in 4.5% of cases (Table 4).

**Table 4.** Analysis of the operators.

|  | Total number<br>(%) | Overall success<br>(%) | First-attempt<br>(%) |
| --- | --- | --- | --- |
| Seniority of the operator |  |  |  |
| - Non-physician: | 19 (1.9) | 19 (100) | 13 (68.4) |
| - Less than 2 y of training: | 373 (37.3) | 363 (97.3) | 313 (83.9) |
| - 2-5 y of training: | 239 (23.9) | 236 (98.7) | 212 (88.7) |
| - More than 5 y of training: | 369 (36.9) | 368 (99.7) | 344 (93.2) |
| Previous experience with the i-gel |  |  |  |
| - None: | 27 (2.7) |  |  |
| - 1-20 devices inserted: | 354 (35.4) |  |  |
| - 20-50 devices inserted: | 171 (17.1) |  |  |
| - More than 50 devices inserted: | 448 (44.8) |  |  |
| i-gel Plus insertion reported as |  |  |  |
| - Very easy: | 443 (44.9) |  |  |
| - Easy: | 349 (35.4) |  |  |
| - Neutral: | 150 (15.2) |  |  |
| - Difficult: | 44 (4.5) |  |  |
| - Very difficult: | 0 |  |  |

Perioperative and postoperative adverse events and patient complaints are summarized in Table 5. Traces of blood on the device at removal were seen in 7.4% of patients, however, none of these required any further intervention. No patient presented with clinical symptoms of gastric content aspiration, and the incidence of perioperative laryngospasm or bronchospasm was extremely low. The most frequently reported postoperative complaint was sore throat, occurring in 12.7% of patients at 24 h. Only 1.4% of these described their sore throat as moderate or severe on a 0-10 scale. A hoarse voice was detected in 4.9% of patients at 24 h, with 0.4% of them being moderate to severe. Fourteen patients (1.4%) reported any numbness inside the oral cavity including the tongue at 24 h. All these patients are further followed for evaluation of the duration and severity of symptoms.

**Table 5.**
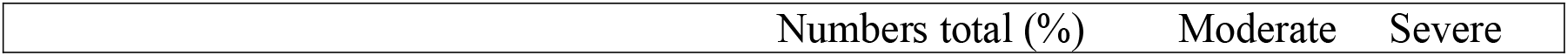
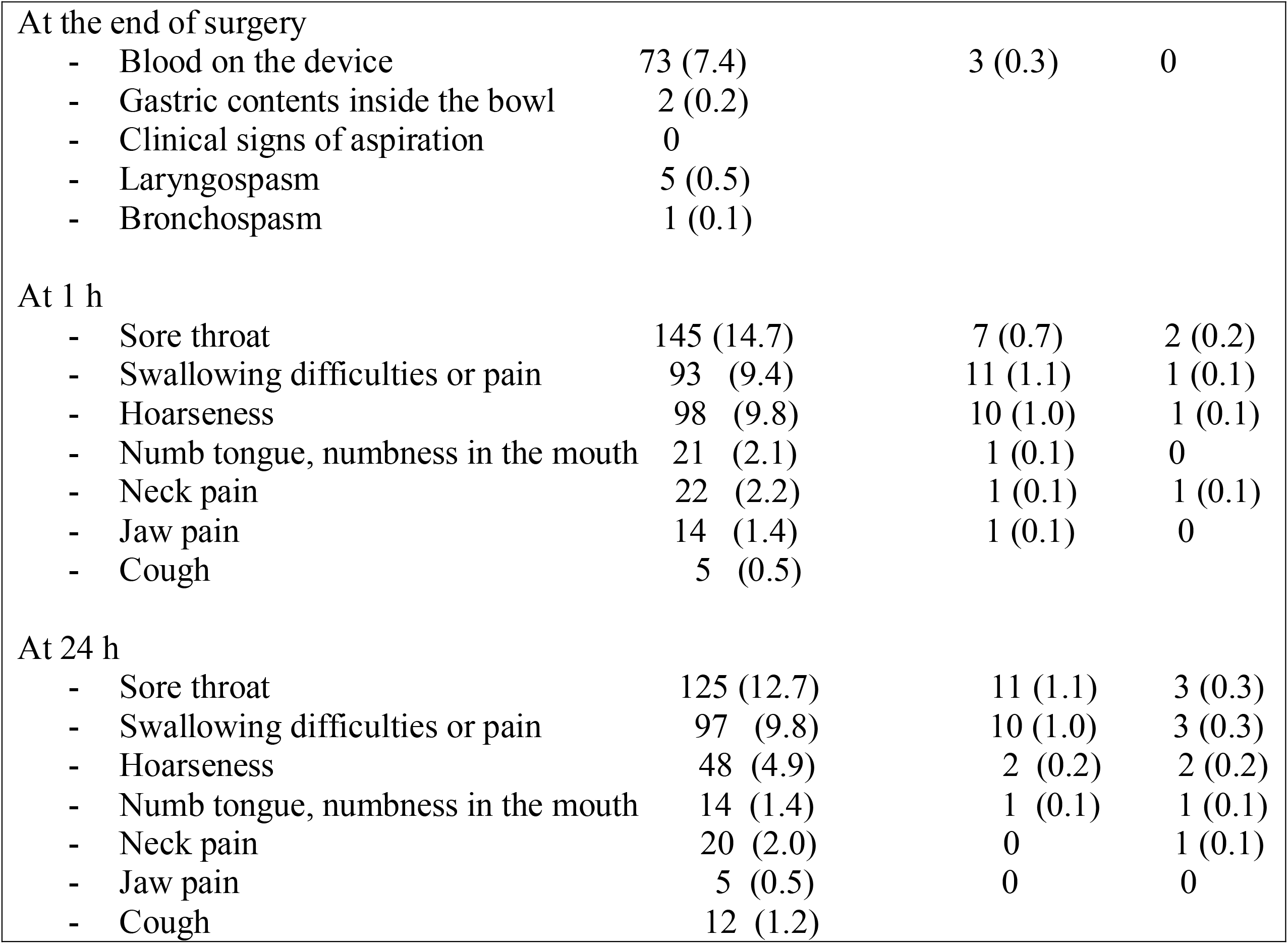
Perioperative and postoperative complications.

|  | Numbers total (%) | Moderate | Severe |
| --- | --- | --- | --- |
| At the end of surgery |  |  |  |
| - Blood on the device | 73 (7.4) | 3 (0.3) | 0 |
| - Gastric contents inside the bowl | 2 (0.2) |  |  |
| - Clinical signs of aspiration | 0 |  |  |
| - Laryngospasm | 5 (0.5) |  |  |
| - Bronchospasm | 1 (0.1) |  |  |
| At 1 h |  |  |  |
| - Sore throat | 145 (14.7) | 7 (0.7) | 2 (0.2) |
| - Swallowing difficulties or pain | 93 (9.4) | 11 (1.1) | 1 (0.1) |
| - Hoarseness | 98 (9.8) | 10 (1.0) | 1 (0.1) |
| - Numb tongue, numbness in the mouth | 21 (2.1) | 1 (0.1) | 0 |
| - Neck pain | 22 (2.2) | 1 (0.1) | 1 (0.1) |
| - Jaw pain | 14 (1.4) | 1 (0.1) | 0 |
| - Cough | 5 (0.5) |  |  |
| At 24 h |  |  |  |
| - Sore throat | 125 (12.7) | 11 (1.1) | 3 (0.3) |
| - Swallowing difficulties or pain | 97 (9.8) | 10 (1.0) | 3 (0.3) |
| - Hoarseness | 48 (4.9) | 2 (0.2) | 2 (0.2) |
| - Numb tongue, numbness in the mouth | 14 (1.4) | 1 (0.1) | 1 (0.1) |
| - Neck pain | 20 (2.0) | 0 | 1 (0.1) |
| - Jaw pain | 5 (0.5) | 0 | 0 |
| - Cough | 12 (1.2) |  |  |

## Discussion

This study followed the recommendations suggested by ADEPT [9] and ADEPT-2 [10] initiatives and, in a large sample, focused on the clinically important primary outcome, the total success rate of the i-gel Plus insertion. The interim results of this trial showed that the total success rate of the device insertion is reasonably high in both males and females. The overall success rate of 98.6% is slightly higher than the results published for the i-gel supraglottic device [11]. Theiler et al. [12] reported the total success rate of 96% in a large multicentric cohort study, while in smaller trials this outcome was measured between 84 and 100% [13-15]. This overall insertion success rate of the i-gel Plus is also comparable or superior to other modern second-generation supraglottic devices such as LMA Supreme, LMA Protector, SaCo VLM, or AuraGain laryngeal mask [16-19].

The first-attempt success rate in our study was 88.2% which is significantly lower than in the aforementioned large trial involving the original version of the i-gel [12] where the authors reported 93.4% success on the initial insertion attempt. This may be explained by the larger size of the i-gel Plus’s bowl and by the fact that all insertions were performed initially without the use of muscle relaxation. However, the performance of the initial insertion was significantly affected by the experience of the anaesthetist – senior operators achieved a 93.3% first-attempt success rate while non-physicians or junior trainees with less than 2 years of experience only 83.2%.

Our team also evaluated independent risk factors for the i-gel Plus failure (Table 4). While this was not possible to perform statistically for the overall success rate due to a low number of subjects, a manual logistic multivariate regression provided the date for the failures on the first attempt. Among all of the parameters studied, only the seniority of the anaesthetist significantly affected the initial first-attempt success rate. This contradicts the findings reported by Theiler et al. [12] where less experienced anaesthetists succeeded more frequently than those with more experience. Other factors such as age, gender, or difficult airway predictors surprisingly did not influence the first-time success rate.

Insertion time was reasonably short in most cases. However, it is quite difficult to compare the device insertion time recorded in our study with those presented by other sources, as the methodology of insertion time measurement varies significantly between studies. Therefore we measured two insertion time intervals, the first one from discontinuation of face-mask ventilation until i-gel Plus placement, and the second one until the appearance of the first capnographic trace on the monitor demonstrating gas exchange.

The importance of the oropharyngeal seal pressure (OSP) is often overestimated. Generally, it does not mean that a device exhibiting higher seal pressure has necessarily better clinical performance. In our experience, all SGAs having lower seal pressures than 20 cmH_2_O may not be sufficient for effective ventilation whereas seal pressures over 25 cmH_2_O provide reliable gas exchange in most patients under controlled ventilation. The mean oropharyngeal seal pressure in our study was over 30 cmH_2_O which is superior to the findings reported in Theiler’s evaluation of the i-gel [12]. The proportion of patients with OSPs lower than 20 cmH_2_O in our study was 4.9% while 80% of them had higher OSPs than 25 cmH_2_O.

Fibreoptic examination of the glottis through the device was performed in almost one quarter of patients. This evaluation is important for confirmation of the correct position of the device, which should face directly the vocal cords and the epiglottis should be located outside of the fibreoptic view. In 90.1% of patients, the fibreoptic view provided an either full or almost full view of the laryngeal inlet. This makes the i-gel Plus a reasonable conduit for subsequent tracheal intubation. Theiler et al. do not report fibreoptic evaluation through the i-gel but reliable data may be retrieved from another trial [20]. The authors reported grade 1 or grade 2 views through the i-gel in 83.5% of patients. In a study on fresh cadavers, the i-gel Plus provided better intubation conditions than the i-gel SGA [21].

The overall incidence of postoperative complaints, such as sore throat and swallowing difficulties may seem relatively high in our cohort [22]. However, we recorded all complaints apart from cough on a 0-10 verbal scale. If only moderate and severe symptoms were counted the incidence is similar or lower than in other studies evaluating the i-gel device [14,23]. This study is also the first one to intentionally evaluate long-term complications associated with the insertion of an SGA [24]. The i-gel may rarely cause moderate oropharyngeal trauma [25] or neuropraxia [26], full results of long-term complications related to the i-gel Plus will become available after the completion of this trial.

Limitations of this trial arise mainly from its non-randomized cohort design. There is no direct comparison of the i-gel Plus performance against the original version of the i-gel, or against other modern SGAs such as the LMA Supreme, LMA Protector, or LM AuraGain. Another limitation is that only one-quarter of the patients had fibreoptic evaluation through the device. The main reason for this was the unavailability of a flexible bronchoscope for all cases and centers, as well as the high financial burden for the trial. Only a small percentage of patients had a gastric tube inserted through the gastric channel of the device. We decided that routine gastric tube insertion represents an additional intervention with potential risk for the patient and furthermore, its role in protection against aspiration of gastric contents remains unclear [27]. The i-gel Plus was not also not used in this study for extended applications, such as prone position surgery, shared airway procedure or in obese patients. Oesophageal seal pressure was also not measured and neither was the depth the device tip was inserted into the oesophagus. Currently the oesophageal seal pressure can be measured only experimentally on cadavers, and the assessment of the oesophageal position of the tip requires either an imaging method or flexible oesophagoscopy. One cadaver study showed very low oseophageal seal pressures associated with the i-gel demonstrating its shallow insertion into the oesophagus [28]. We suspect that the longer tip of the i-gel Plus device would provide significantly higher oesophageal seal pressures than those found in the original device, but this requires further study.

Based on these interim results we conclude that the i-gel Plus SGA is a promising device. In this initial evaluation of our study results, it exhibited a high overall success rate of insertion and achieved oropharyngeal seal pressures which enabled effective ventilation and oxygenation in the vast majority of patients. Its use is associated with a low incidence of serious complications. Evaluation of this device as a conduit for tracheal intubation, in resuscitation scenarios, and for other indications should be performed in the near future.

## Supporting information

Study supplement 1, Case report form

## Data Availability

All data produced in the present study are available upon reasonable request to the authors

## Acknowledgements

We thank all study investigators not listed as the authors: M. Lukes, J. Janouskova, M. Mlejnecka, K. Kapralova, T. K. Bhoday, J. Ulrichova, J. Blaha, T. Brozek, M. Kral, L. Gomez, J. Holland, F. McAleavey, L. Merjava Skripecka, S. Sobczyk, J. Maiboroda, L. Sindelar, D. Novotny, K. Subrtova.

## Funding Statement

This trial is supported by the Czech Ministry of Health, grant no. MZCZ-DRO-VFN64165.

The details of the Ethical Committees/IRBs that provided approval for this trial are given below:

Ethics Committee of the General University Hospital in Prague (study sponsor) in November 2019 (No. 1952/19 S-IV, received 14/11/2019, dr. J. Sedivy, Chair). The Czech Ministry of Health will act as a sponsor and partial founder via General University Hospital in Prague (grant number MZCZ-DRO-VFN64165). Following this initial approval, the ethical approvals from other centres were obtained: University Hospital Olomouc (No. 18/20, received 10/02/2020, Dr. J. Buresova, chair), University Hospital in Lodz (No. RNN/61/20/KE, received 03/03/2020, Prof. J. Drzewoski, chair), University Hospital in Barcelona (No. HCB 2020/0771, received 08/09/2020, Dr. A.L. Arellano Andrino, secretary), Northern HSC Trust for Antrim Area Hospital in Antrim (No. NT20-278410-10, received 10/12/2020, Dr. M. Rooney, Trust Director of Research and Development), Southern HSC Trust for Craigavon Area Hospital (No. ST2021/31, received 22/04/2021, Dr. P. Sharpe, Trust Director of Research and Development), Office for Research and Ethical Committee Northern Ireland (ORECNI) (REC 20/NI/0140, received 27/11/2020, Prof. P. Murphy, HSC REC B Chair), University Military Hospital in Prague (No. 108/15-104/2020, received 21/12/2020, Prof. J. Plas, chair).

