## Supplementary material for "An evaluation of the i-gel Plus® supraglottic airway device in elective patients: the interim results from a prospective international multicentre study": Study supplement 1, Case report form

### Patient Data

Record ID ______________________

Hospital
- ◯ ES / Barcelona
- ◯ CZ / GUH Prague
- ◯ CZ / GUH Olomouc
- ◯ PL / MUL
- ◯ UK / AAH
- ◯ UK / CAH
- ◯ CZ / MUH

#### Enrollment

Documents:

#### Inclusion Criteria

| | Yes | No |
|---|---|---|
| Age 18-89 years | ◯ | ◯ |
| ASA status I, II or III | ◯ | ◯ |
| Elective procedure without need for muscle relaxation | ◯ | ◯ |
| Understanding the "Information for the patients" leaflet | ◯ | ◯ |

#### Exlusion Criteria

| | Yes | No |
|---|---|---|
| Age less than 18 or more than 89 years | ◯ | ◯ |
| Emergency surgery | ◯ | ◯ |
| Intraabdominal operations | ◯ | ◯ |
| Intrathoracic procedures | ◯ | ◯ |
| Shared airway surgery | ◯ | ◯ |
| Increased risk for aspiration of gastric contents | ◯ | ◯ |
| BMI of more than 35 kg/m2 | ◯ | ◯ |
| Unusual operating positioning - steep head down, prone, sitting | ◯ | ◯ |
| Incapacity to understand/sign informed consent (learning difficulties, language difficulties). | ◯ | ◯ |

Informed consent signed:
- ◯ Yes
- ◯ No

Informed consent date/time: ______________________

| | |
|---|---|
| Enrolled: | ◯ Yes<br>◯ No |
| Enrolled date/time: | __________________ |

#### Demographic data

| | |
|---|---|
| Age: | __________________ |
| Gender: | ◯ Male<br>◯ Female |
| Height [cm] | __________________ |
| Weight [kg] | __________________ |
| BMI | __________________ |

#### Preoperative

| | |
|---|---|
| ASA clasification: | ◯ 1<br>◯ 2<br>◯ 3<br>◯ 4 |

#### Associated conditions

| | |
|---|---|
| Ischaemic heart disease: | ◯ Yes<br>◯ No |
| Hypertension: | ◯ Yes<br>◯ No |
| Cerebrovascular disease: | ◯ Yes<br>◯ No |
| COPD: | ◯ Yes<br>◯ No |
| Asthma: | ◯ Yes<br>◯ No |
| Diabetes: | ◯ Yes<br>◯ No |
| Liver disease: | ◯ Yes<br>◯ No |
| Kidney disease: | ◯ Yes<br>◯ No |

Other: ______________________

Allergy: ◯ Yes ◯ No

Alergy info ______________________

#### Airway Assessment

Mallampati: ◯ 1 ◯ 2 ◯ 3 ◯ 4

Wilson grade: ◯ A ◯ B ◯ C

Thyromental distance [cm]: ______________________

Neck circumference [cm] ______________________

Neck movement: ◯ good ◯ limited ◯ minimal

Mouth opening [cm]: ______________________

Dentition: ◯ Sound ◯ Crowns/bridges/venirs ◯ Dentures/toothless

Beard: ◯ Yes ◯ No

#### Intraoperative

#### Procedure

Procedure type: ◯ General surgery ◯ Gynaecology ◯ Urology ◯ Vascular surgery ◯ Orthopaedics / trauma ◯ Other

Name of procedure: ______________________

| | |
|---|---|
| Duration [min.]: | __________________________________ |
| Position: | ◯ Supine<br>◯ Lateral<br>◯ Gynaecology<br>◯ Trendelenburg<br>◯ anti-Trendelenburg<br>◯ Other |
| Operator's experience:<br>Number of previous i-gel (original version) insertions | ◯ 0<br>◯ 1-20<br>◯ 21-50<br>◯ 51+ |
| Operator`s seniority | ◯ Non-physician<br>◯ Less than 2 years of training<br>◯ 2-5 years of training<br>◯ More than 5 years of training / specialty doctor / consultant |
| Device size: | ◯ 3<br>◯ 4<br>◯ 5 |

#### Induction to anaesthesia

| | |
|---|---|
| Propofol [mg]: | __________________________________ |
| Other induction agent than Propofol [mg] | __________________________________ |
| Opioid: | ◯ Fentanyl<br>◯ Sufentanil<br>◯ Alfentanil<br>◯ Remifentanil<br>◯ Morphine |
| Opioid dose: | __________________________________ |
| Opioid units | ◯ mcg<br>◯ mg |

#### Maintenance

| | |
|---|---|
| Muscle relaxant:<br>(it is not a part of the protocol. If given, for any reason, this must be reported to the CRF). | __________________________________ |
| Inhalation anesthetics | ◯ Sevoflurane<br>◯ Desflurane<br>◯ None-TIVA |

| | |
|---|---|
| Opioid: | ◯ Fentanyl<br>◯ Sufentanil<br>◯ Alfentanil<br>◯ Remifentanil<br>◯ Morphine |
| Total opioid dose: | ______________________ |
| Opioid units | ◯ mcg<br>◯ mg |
| Non-opioid analgesics - drug name: | ______________________ |
| Non-opioid analgesics drug dose [mg] | ______________________ |
| 2nd Non-opioid analgesics drug name: | ______________________ |
| 2nd Non-opioid analgesics drug dose [mg] | ______________________ |
| 3rd Non-opioid analgesics drugs name: | ______________________ |
| 3rd Non-opioid analgesics drug dose [mg] | ______________________ |

#### Respiratory

| | |
|---|---|
| Bag - mask ventilation | ◯ Easy<br>◯ Easy two persons<br>◯ Easy / Guedelairway<br>◯ Difficult<br>◯ Impossible |
| Ventilation | ◯ VCV<br>◯ PCV<br>◯ SIMV<br>◯ PSV<br>◯ Spontaneous |
| Monitoring | ☐ ECG<br>☐ NIPB<br>☐ Pulse oximetry<br>☐ Capnography<br>☐ Invasive arterial pressure<br>☐ CVP |
| Peak pressure max. [cm H2O] | ______________________ |
| Compliance [ml * cm H2O-1]: | ______________________ |

| | |
|---|---|
| Inhalation gas: | ◯ Air / Oxygen<br>◯ Oxygen / Nitrous |

#### Outcomes

#### Primary outcome

| | |
|---|---|
| Success: | ◯ Yes<br>◯ No |
| Number of attempts: | ◯ 1<br>◯ 2<br>◯ 3 |
| In case of failure: | ◯ Tracheal intubation<br>◯ Other SGA |
| Other SGA (name) | ______________________ |
| Different size of the i-gel Plus | ◯ Yes<br>◯ No |
| What size of i-gel was used? | ◯ 3<br>◯ 4<br>◯ 5 |

#### Secondary outcomes

Total gas flow is set at 5 l.min-1, the Adjustable Pressure Limiting (APL) valve is set to 40 cmH2O. The operator waits until an audible leak is detected from around the device – the stethoscope placed to the jugulum. The pressure at which the leak is noted is recorded. If the pressure reaches 40 cmH2O before a leak is detected the leak pressure is recorded as 40 cmH2O. The pressure gauge on the anaesthetic machine is used to record the pressure.

| | |
|---|---|
| Seal (leak) pressure after the insersion [cm H2O]:<br>(maximum 40 cmH2O) | ______________________ |
| Seal (leak) pressure at 30 min [ cm H2O]: | ______________________ |
| Seal (leak) pressure before the end [ cm H2O]: | ______________________ |
| Insertion time 1 [sec]: | ______________________<br>(From grasping the i-gel to placement) |
| Insertion time 2 [sec.] | ______________________<br>(From grasping the i-gel to the first etCO2) |

| | |
|---|---|
| Lowest spO2 during induction (%) | ________________ (0-100) |
| i-Gel Plus intraoperative manipulation required | ◯ Yes<br>◯ No |
| Change of the position | ☐ Change of the device position<br>☐ Head or neck positioning<br>☐ Jaw thrust |
| Subjective assessment of insertion ease: | ◯ Very easy<br>◯ Easy<br>◯ Neutral<br>◯ Difficult<br>◯ Very difficult |
| Fiberoptic assessment of the device position: | ◯ Yes<br>◯ No |
| Device position: | ◯ 1<br>◯ 2<br>◯ 3<br>◯ 4 |
| Insertion of the gastric tube: | ◯ Yes<br>◯ No |
| Gastric tube insersion ease: | ◯ Very easy<br>◯ Easy<br>◯ Neutral<br>◯ Difficult<br>◯ Very difficult |

**At the end of surgery**

| | |
|---|---|
| Blood on the device: | ◯ Yes<br>◯ No |
| Gastric contents inside the bowl: | ◯ Yes<br>◯ No |
| Clinical signs of aspiration: | ◯ Yes<br>◯ No |
| Laryngospasm: | ◯ Yes<br>◯ No |
| Bronchospasm: | ◯ Yes<br>◯ No |
| Other: | ________________ |

#### Postoperative

##### Assesment 1 h post awakening

| | 0 | 1 | 2 | 3 | 4 | 5 | 6 | 7 | 8 | 9 | 10 |
|---|---|---|---|---|---|---|---|---|---|---|---|
| Sore throat (0-10 scale): | ◯ | ◯ | ◯ | ◯ | ◯ | ◯ | ◯ | ◯ | ◯ | ◯ | ◯ |
| Pain on swallowing, swallowing difficulties (0-10 scale): | ◯ | ◯ | ◯ | ◯ | ◯ | ◯ | ◯ | ◯ | ◯ | ◯ | ◯ |
| Hoarseness (0-10 scale): | ◯ | ◯ | ◯ | ◯ | ◯ | ◯ | ◯ | ◯ | ◯ | ◯ | ◯ |
| Numb tongue, numbness inside the oral cavity (0-10 scale): | ◯ | ◯ | ◯ | ◯ | ◯ | ◯ | ◯ | ◯ | ◯ | ◯ | ◯ |
| Neck pain (0-10 scale): | ◯ | ◯ | ◯ | ◯ | ◯ | ◯ | ◯ | ◯ | ◯ | ◯ | ◯ |
| Jaw pain (0-10 scale): | ◯ | ◯ | ◯ | ◯ | ◯ | ◯ | ◯ | ◯ | ◯ | ◯ | ◯ |

Cough:
◯ Present preoperatively
◯ Yes
◯ No

Cough type:
◯ Dry
◯ Wet

##### Assessment at 24 h post awakening

| | 0 | 1 | 2 | 3 | 4 | 5 | 6 | 7 | 8 | 9 | 10 |
|---|---|---|---|---|---|---|---|---|---|---|---|
| Sore throat (0-10 scale): | ◯ | ◯ | ◯ | ◯ | ◯ | ◯ | ◯ | ◯ | ◯ | ◯ | ◯ |
| Pain on swallowing, swallowing difficulties (0-10 scale): | ◯ | ◯ | ◯ | ◯ | ◯ | ◯ | ◯ | ◯ | ◯ | ◯ | ◯ |
| Hoarseness (0-10 scale): | ◯ | ◯ | ◯ | ◯ | ◯ | ◯ | ◯ | ◯ | ◯ | ◯ | ◯ |
| Numb tongue, numbness inside the oral cavity (0-10 scale): | ◯ | ◯ | ◯ | ◯ | ◯ | ◯ | ◯ | ◯ | ◯ | ◯ | ◯ |
| Neck pain (0-10 scale): | ◯ | ◯ | ◯ | ◯ | ◯ | ◯ | ◯ | ◯ | ◯ | ◯ | ◯ |
| Jaw pain (0-10 scale): | ◯ | ◯ | ◯ | ◯ | ◯ | ◯ | ◯ | ◯ | ◯ | ◯ | ◯ |

Cough:
◯ Present preoperatively
◯ Yes
◯ No

Cough type:
◯ Dry
◯ Wet

#### Long term follow up at 3 month (TELEPHONE REVIEW)

Only in patients reporting sore throat, pain on swallowing, neck pain or jaw pain more than 5 (on 0-10 scale) at 24 h
In all patients reporting hoarseness or numb tongue/numbness inside the oral cavity at 24 h

| | |
|---|---|
| Symptoms persisting | ◯ Yes<br>◯ No |
| Sore throat | ◯ Yes<br>◯ No |
| Pain on swallowing, swallowing difficulties: | ◯ Yes<br>◯ No |
| Hoarseness: | ◯ Yes<br>◯ No |

All patients reporting long term hoarseness (which was not present preoperatively) should have endoscopic or other assessment of vocal cord mobility.

| | |
|---|---|
| Free text:<br>Comments on whether the vocal cords hypokinesis or paralysis is present:<br>E. g.: "left vocal cord completely immobile, in open position". | ______________________________ |
| Numb tongue, numbness inside the oral cavity: | ◯ Yes<br>◯ No |
| Free text:<br>Comments on localization of numbness/motor deficit, e. g. "numb left half of the tongue" or "numb lower lip". | ______________________________ |
| Neck pain: | ◯ Yes<br>◯ No |
| Jaw pain: | ◯ Yes<br>◯ No |

#### Long term follow up at 6 month (TELEPHONE REVIEW)

Only in patients reporting sore throat, pain on swallowing, neck pain or jaw pain more than 5 (on 0-10 scale) at 24 h
In all patients reporting hoarseness or numb tongue/numbness inside the oral cavity at 24 h

| | |
|---|---|
| Symptoms persisting | ◯ Yes<br>◯ No |
| Sore throat | ◯ Yes<br>◯ No |
| Pain on swallowing, swallowing difficulties: | ◯ Yes<br>◯ No |

| | |
|---|---|
| Hoarseness: | ◯ Yes<br>◯ No |
| All patients reporting long term hoarseness (which was not present preoperatively) should have endoscopic or other assessment of vocal cord mobility. | |
| Free text:<br>Comments on whether the vocal cords hypokinesis or paralysis is present:<br>E. g.: "left vocal cord completely immobile, in open position". | __________________________ |
| Numb tongue, numbness inside the oral cavity: | ◯ Yes<br>◯ No |
| Free text:<br>Comments on localization of numbness/motor deficit, e. g. "numb left half of the tongue" or "numb lower lip". | __________________________ |
| Neck pain: | ◯ Yes<br>◯ No |
| Jaw pain: | ◯ Yes<br>◯ No |
